# Concurrent validity and reliability of suicide risk assessment instruments: A meta analysis of 20 instruments across 27 international cohorts

**DOI:** 10.1101/2021.09.15.21263562

**Authors:** Adrian I. Campos, Laura S. Van Velzen, Dick J. Veltman, Elena Pozzi, Sonia Ambrogi, Elizabeth D. Ballard, Nerisa Banaj, Zeynep Başgöze, Sophie Bellow, Francesco Benedetti, Irene Bollettini, Katharina Brosch, Erick J. Canales-Rodríguez, Emily K. Clarke-Rubright, Lejla Colic, Colm G. Connolly, Philippe Courtet, Kathryn R. Cullen, Udo Dannlowski, Maria R. Dauvermann, Christopher G. Davey, Jeremy Deverdun, Katharina Dohm, Tracy Erwin-Grabner, Negar Fani, Lydia Fortea, Paola Fuentes-Claramonte, Ali Saffet Gonul, Ian H. Gotlib, Dominik Grotegerd, Mathew A. Harris, Ben J. Harrison, Courtney C. Haswell, Emma L. Hawkins, Dawson Hill, Yoshiyuki Hirano, Tiffany C. Ho, Fabrice Jollant, Tanja Jovanovic, Tilo Kircher, Bonnie Klimes-Dougan, Emmanuelle le Bars, Christine Lochner, Andrew M. McIntosh, Susanne Meinert, Yara Mekawi, Elisa Melloni, Philip Mitchell, Rajendra A. Morey, Akiko Nakagawa, Igor Nenadić, Emilie Olié, Fabricio Pereira, Rachel D. Phillips, Fabrizio Piras, Sara Poletti, Edith Pomarol-Clotet, Joaquim Radua, Kerry J. Ressler, Gloria Roberts, Elena Rodriguez-Cano, Matthew D. Sacchet, Raymond Salvador, Anca-Larisa Sandu, Eiji Shimizu, Aditya Singh, Gianfranco Spalletta, J. Douglas Steele, Dan J. Stein, Frederike Stein, Jennifer S. Stevens, Giana I. Teresi, Aslihan Uyar-Demir, Nic J. van der Wee, Steven J. van der Werff, Sanne J.H. van Rooij, Daniela Vecchio, Norma Verdolini, Eduard Vieta, Gordon D. Waiter, Heather Whalley, Sarah L. Whittle, Tony T. Yang, Carlos A. Zarate, Paul M. Thompson, Neda Jahanshad, Anne-Laura van Harmelen, Hilary P. Blumberg, Lianne Schmaal, Miguel E. Rentería

## Abstract

**Objective:** A major limitation of current suicide research is the lack of power to identify robust correlates of suicidal thoughts or behaviour. Variation in suicide risk assessment instruments used across cohorts may represent a limitation to pooling data in international consortia.

**Method:** Here, we examine this issue through two approaches: (i) an extensive literature search on the reliability and concurrent validity of the most commonly used instruments; and (ii) by pooling data (N∼6,000 participants) from cohorts from the ENIGMA-Major Depressive Disorder (ENIGMA-MDD) and ENIGMA-Suicidal Thoughts and Behaviour (ENIGMA-STB) working groups, to assess the concurrent validity of instruments currently used for assessing suicidal thoughts or behaviour.

**Results:** Our results suggested a pattern of moderate-to-high correlations between instruments, consistent with the wide range of correlations, r=0.22-0.97, reported in the literature. Two common complex instruments, the Columbia Suicide Severity Rating Scale (C-SSRS) and the Beck Scale for Suicidal Ideation (SSI), were highly correlated with each other (r=0.83), as were suicidal ideation items from common depression severity questionnaires.

**Conclusions:** Our findings suggest that multi-item instruments provide valuable information on different aspects of suicidal thoughts or behaviour, but share a core factor with single suicidal ideation items found in depression severity questionnaires. Multi-site collaborations including cohorts that used distinct instruments for suicide risk assessment should be feasible provided that they harmonise across instruments or focus on specific constructs of suicidal thoughts or behaviours.

**Key points:**

- Question: To inform future suicide research in multi-site international consortia, it is important to examine how different suicide measures relate to each other and whether they can be used interchangeably.
- Findings: Findings suggest detailed instruments (such as the Columbia Suicide Severity Rating Scale and Beck Scale for Suicidal Ideation) provide valuable information on suicidal thoughts and behaviour, and share a core factor with items on suicidal ideation from depression severity rating scale (such as the Hamilton Depression Rating Scale or the Beck Depression Inventory).
- Importance: Results from international collaborations can mitigate biases by harmonising distinct suicide risk assessment instruments.
- Next steps: Pooling data within international suicide research consortia may reveal novel clinical, biological and cognitive correlates of suicidal thoughts and/or behaviour.

## Introduction

Suicide is a leading cause of death worldwide, with an estimated 800,000 deaths by suicide occurring annually, or one person dying by suicide every 40 seconds (World Health Organization, 2014). Despite national and international efforts to prevent suicide, suicide rates continue to rise around the world (Alicandro et al., 2019).

To better understand and identify demographic, environmental, psychological, cognitive and neurobiological factors associated with suicidal thoughts and behaviour, we need large samples, as individual factors most likely explain a small proportion of complex phenotypes as suicidal thoughts or behaviours. Large and diverse samples additionally provide the opportunity to study the heterogeneity in associated factors by identifying subgroups. Large-scale international collaborations in consortia for suicide research may provide an important step forward.

One example of these consortia is the Enhancing NeuroImaging Genetics through Meta-Analysis Suicidal Thoughts and Behaviour (ENIGMA-STB) consortium. The aim of ENIGMA-STB is to study the neural correlates of suicidal thoughts and behaviour (STB), by bringing together research groups around the world that have collected both neuroimaging data and assessed STB in individuals with and without mental disorders. These groups use standardized protocols to process their neuroimaging data and then pool data in analyses that have increased statistical power to detect relevant associations.

While these large-scale collaborations have many strengths, it has been challenging to harmonize scores from the different instruments employed to assess STBs across cohorts, and the validity of the findings will depend on the quality of STB measure harmonization. For instance, in our recent large-scale analysis of cortical morphology across 18 research groups within the ENIGMA Major Depressive Disorder (ENIGMA-MDD) consortium (Campos et al., 2020), STBs were assessed using 19 different measures, including single items on STBs from depression severity questionnaires, items from clinical interviews, in addition to items from comprehensive instruments specifically focused on STBs such as the Columbia Suicide Severity Rating Scale (C-SSRS) (Posner et al., 2008; Posner et al., 2011).

To inform future suicide research in international consortia, it is important to examine how these different suicide measures relate to each other and whether they can be used interchangeably. Therefore, the aim of this study was to examine the correlations between the 20 different assessment instruments for STBs used across 27 ENIGMA cohorts. In the first part of this report, we provide an overview of the literature on reliability and validity of commonly-used measures to assess STBs, and the associations between these measures (concurrent validity). In the second part, we present findings from a meta-analysis performed using the measures collected across 27 cohorts within the ENIGMA-MDD and ENIGMA-STB working groups.

## Methods

### Literature search

A literature search was conducted in PubMed (https://pubmed.ncbi.nlm.nih.gov) for articles published before September 2020, using the following search terms: suicid* AND (questionnaire* OR interview OR measures) AND (validity OR convergent validity OR discriminant validity OR reliability OR psychometric*), using ‘English’ and ‘Human’ as additional filters.

1,156 abstracts were screened by investigator LvV to identify studies which used psychometric measures to assess suicidal ideation and/or suicidal behaviour that were also collected by the ENIGMA research groups. These measures included: the Beck Depression Inventory (BDI) suicidal ideation item (Beck et al., 1961, 1996), Scale for Suicidal Ideation (SSI) (Beck et al., 1988; Beck et al., 1979), Children’s Depression Rating Scale (CDRS) suicidal ideation item (Poznanski & Mokros, 1996), Composite International Diagnostic Interview (CIDI) items on suicidal ideation and behaviour (WHO, 1997), Columbia Suicide Severity Rating Scale (C-SSRS) (Posner et al., 2011), Diagnostic Interview for Genetics Studies (DIGS) items on suicidal ideation and behaviour (Nurnberger et al., 1994), Hamilton Depression Rating Scale (HAM-D) item on suicidal ideation (Hamilton, 1960), Inventory of Depression and Anxiety Symptoms (IDAS-II) suicide subscale (Watson et al., 2012), (Quick) Inventory of Depressive Symptomatology (IDS/QIDS) suicidal ideation item (Rush et al., 1986; Rush et al., 2003), Kiddie Schedule for Affective Disorders and Schizophrenia (KSADS) suicide items (Kaufman et al., 1997), Montgomery-Asberg Depression Rating Scale (MADRS) suicidal ideation item (Montgomery & Asberg, 1979), Mini International Neuropsychiatric Interview (MINI) suicidality module (Sheehan et al., 1998), Revised Children’s Anxiety and Depression Scale (RCADS) suicidal ideation item (Chorpita et al., 2000), Structured Clinical Interview for DSM Disorders (SCID) suicide questions (First, 1997), Suicidal Ideation Questionnaire (SIQ) (Reynolds, 1987), Beck’s Suicide Intent Scale (SIS) (Beck et al., 1974), Self-Injurious Thoughts and Behaviours Interview (SITBI) (Nock et al., 2007), Suicide Score Scale (SSS) (Innamorati et al., 2008), Youth Self-Report suicide item (YSR) (Achenbach et al., 1991), Suicidal Ideation Questionnaire-Junior (SIQ-JR) (Reynolds, 1987).

A total of 180 studies were identified and screened for information on the reliability (inter-rater reliability, internal consistency or test-retest reliability) or validity (correlation with an established instrument e.g., concurrent validity or predictive validity) of those measures. For concurrent validity, we included only associations between measures that were collected by the ENIGMA working groups and mentioned above. Additional studies were identified by cross-referencing.

### Data dimensionality reduction strategy

Our study comprised both complex (multiple-item) and single-item suicide risk assessment instruments. Single-item instruments, such as questions from depression severity rating scales normally assess recent suicidal ideation. Complex instruments typically capture other dimensions such as control over suicidal thoughts, protective factors and, in the case of suicide attempt, degree of intent to die. By extracting common factor scores for the complex instruments, we are able to obtain a score of the underlying suicidal liability being measured by the instruments while reducing the need to adjust for slightly different wording between versions. The choice of dimensionality reduction approach, common factor scores using full-information maximum likelihood (FIML), was motivated by two reasons; (i) this approach deals with missing data, which is common in these questionnaires, using FIML and (ii) we obtain a single factor score capturing the main liability measured by the instrument, as opposed to other approaches (e.g., PCA) that require non-missing data and output several new variables. Data dimensionality reduction for the SSI, KSADS, IDAS, SIQ, DIGS, CSSRS, MINI and SSS was performed by extracting common factor scores using FIML factor analysis as implemented in the structural equation modelling package *umx* in R (Bates, 2018; Bates et al., 2016).

### Main analysis on ENIGMA data

We implemented a sample size-weighted meta-analysis of correlations between suicide-risk assessment instruments across 27 international cohorts from the ENIGMA Major Depressive Disorder (MDD) and Suicidal Thoughts and Behaviour (STB) working groups. Cohorts shared de-identified individual level response data to STBs assessment instruments or items on STBs from depression symptom severity questionnaires or clinical interviews. All participants provided informed consent and all projects were approved by their respective relevant ethics committees. Our initial analysis consisted of three steps: i) data dimensionality reduction for complex suicide risk assessment instruments (see above); ii) within-cohort unadjusted correlations for all possible pairs of instruments; and iii) a sample-size weighted meta-analysis, averaging the correlation coefficients for pairs of instruments for which data was available across multiple cohorts. Within-cohort correlations, and the sample-size weighted meta-analysis were calculated in python using the *scipy* (Virtanen et al., 2020), *numpy* (Harris et al., 2020) and *pandas* (McKinney & Others, 2010) libraries. Data was visualised using undirected graphs with varying node and edge sizes according to the number of cohorts and cohort pairs supporting each correlation. These were generated from the data using python and the *networkx* library (Hagberg et al., 2008). We analysed data from two working groups of the ENIGMA consortium, including 22 instruments across 27 cohorts worldwide. Individual level responses for 6,716 participants were included in our study (**Table 1**). Participants were included across a range of diagnoses including: major depressive disorder, psychotic disorders, anxiety disorders, obsessive-compulsive disorders, posttraumatic stress disorder, and bipolar disorder, along with data from healthy controls.

**Table 1.** Cohorts included, sample size and instruments used to assess suicidal thoughts and/or behaviours

| Cohort name | Sample size | Instruments |
| --- | --- | --- |
| AFFDIS | 29 | Hamilton Depression Rating Scale (past week), Beck Depression Inventory (past two weeks), Montgomery-Asberg Depression Rating Scale (past week) |
| Chiba University | 117 | Beck Depression Inventory-I (past week), Beck Depression Inventory-II (past two weeks), Mini International Neuropsychiatric Interview suicidality module (past month, question 6 refers to lifetime attempt) |
| Duke/Durham VA | 190 | Beck Depression Inventory (past two weeks), Beck Scale for Suicidal Ideation (past week) |
| EPISCA (Leiden adolescents) | 71 | Children's Depression Inventory (past two weeks), Youth Self-report (lifetime), Revised Children's Anxiety and Depression Scale (lifetime) |
| ETPB-STB | 60 | Hamilton Depression Rating Scale (past week), Scale for Suicidal Ideation (past week), Beck Depression Inventory (past week), Montgomery-Asberg Depression Rating Scale (past week) |
| FIDMAG | 284 | Hamilton Depression Rating Scale (past week), Montgomery-Asberg Depression Rating Scale (past week) |
| FOR2107 Muenster | 424 | Hamilton Depression Rating Scale (past week), Beck Depression Inventory (past week) |
| FOR2107 Marburg | 792 | Hamilton Depression Rating Scale (past week), Beck Depression Inventory (past week) |
| Grady Trauma Project Emory University | 123 | Beck Depression Inventory (past two weeks), Columbia Suicide Severity Rating Scale (past month) |
| The University of Melbourne | 283 | Quick Inventory of Depressive Symptoms (past week), Suicidal Ideation Questionnaire (lifetime), Columbia Suicide Severity Rating Scale (past month and lifetime), Montgomery-Asberg Depression Rating Scale (past week) |
| University of Minnesota Adolescent MDD | 110 | Children's Depression Rating Scale (past week), Beck Depression Inventory (past two weeks), Kiddie Schedule for Affective Disorders and Schizophrenia (lifetime), Inventory of Depression and Anxiety Symptoms (past two weeks) |
| CHU Montpellier BICS study | 66 | Inventory of Depressive Symptoms - Clinician rated (past week), Beck Depression Inventory (current), Columbia Suicide Severity Rating Scale (lifetime & last month) |
| CHU Montpellier IMPACT study | 40 | Quick Inventory of Depressive Symptoms (past week), Inventory of Depressive Symptoms - Clinician rated (past week), Columbia Suicide Severity Rating Scale (last month) |
| CHU Montpellier Servier Study | 120 | Hamilton Depression Rating Scale (past week), Beck Depression Inventory (past week), Scale for Suicidal Ideation (past week) |
| McGill University | 103 | Beck Depression Inventory (past two weeks), Suicide Intent Scale (most recent attempt and most severe attempt), Scale for Suicidal Ideation (today and in the past two weeks), Quick Inventory of Depressive Symptoms (past week), Montgomery-Asberg Depression Rating Scale |
|  |  | (past week), Hamilton Depression Rating Scale (past week), Columbia Suicide Severity Rating Scale (past month and lifetime) |
| Moral Dilemma | 62 | Quick Inventory of Depressive Symptoms (past week), Montgomery-Asberg Depression Rating Scale (past week) |
| Muenster Neuroimaging Cohort | 1064 | Hamilton Depression Rating Scale (past week), Beck Depression Inventory (past week), Montgomery-Asberg Depression Rating Scale (past week) |
| Fondazione Santa Lucia | 288 | Suicide Score Scale (past year), Mini International Neuropsychiatric Interview suicidality module (past month) |
| San Raffaele Hospital | 447 | Hamilton Depression Rating Scale (past week), Beck Depression Inventory (past two weeks), Beck Scale for Suicidal Ideation (day of assessment) |
| SoCAT | 179 | Hamilton Depression Inventory (past week), Beck Depression Inventory (past two weeks) |
| South Africa | 117 | Diagnostic Interview for Genetic Studies (lifetime), Montgomery-Asberg Depression Rating Scale (past week) |
| Stanford University adolescent MDD TIGER | 49 | Columbia Suicide Severity Rating Scale (lifetime & current (ideation in the past week, attempt in the past month), Self-Injurious Thoughts and Behaviours Interview (lifetime) |
| Stanford University AGG/FAA | 56 | Structured Clinical Interview for DSM Disorders (lifetime), Hamilton Depression Rating Scale (past week) |
| STRADL | 1188 | Structured Clinical Interview for DSM Disorders (past month), Quick Inventory of Depressive Symptoms (past week) |
| Sydney Bipolar Risk Study | 225 | Kiddie Schedule for Affective Disorders and Schizophrenia (lifetime), Diagnostic Interview for Genetic Studies (lifetime), Montgomery-Asberg Depression Rating Scale (lifetime) |
| UCSF Adolescent MDD | 161 | Beck Depression Inventory (two weeks), Columbia Suicide Severity Scale (lifetime and current (ideation in the past week, attempt in the past two weeks)) |
| Yale School of Medicine | 178 | Beck Scale for Suicidal Ideation (past month and lifetime), Suicide Intent Scale (most recent attempt, most lethal attempt), Columbia Suicide Severity Rating Scale (lifetime & past month), Hamilton Depression Rating Scale (past week), Child Depression Rating Scale (past week) |

## Results

### Literature review

An overview of the reliability and validity measures for the different suicide scales and items derived from our literature review are presented in **Supplementary Table S1** and **S2**, respectively. Information on the reliability (inter-rater reliability, internal consistency and test-retest reliability) and validity (concurrent and predictive validity) of these measures was most often available for instruments specifically focused on STBs (e.g., SIS, SSI, C-SSRS), followed by items on suicidal ideation from questionnaires assessing severity of depressive symptoms (e.g., BDI, MADRS, HAM-D). No reliability or validity measures were identified for suicide questions from diagnostic interviews (e.g., CIDI, SCID). The lowest concurrent validity measure identified was between the SIS and C-SSRS scale. Overall, mostly high to moderate correlation or concurrent validity scores (kappa range: 0.3-0.97; *r* range: 0.22-0.94) between instruments were identified (**Figure 1a)**.

**Figure 1.**
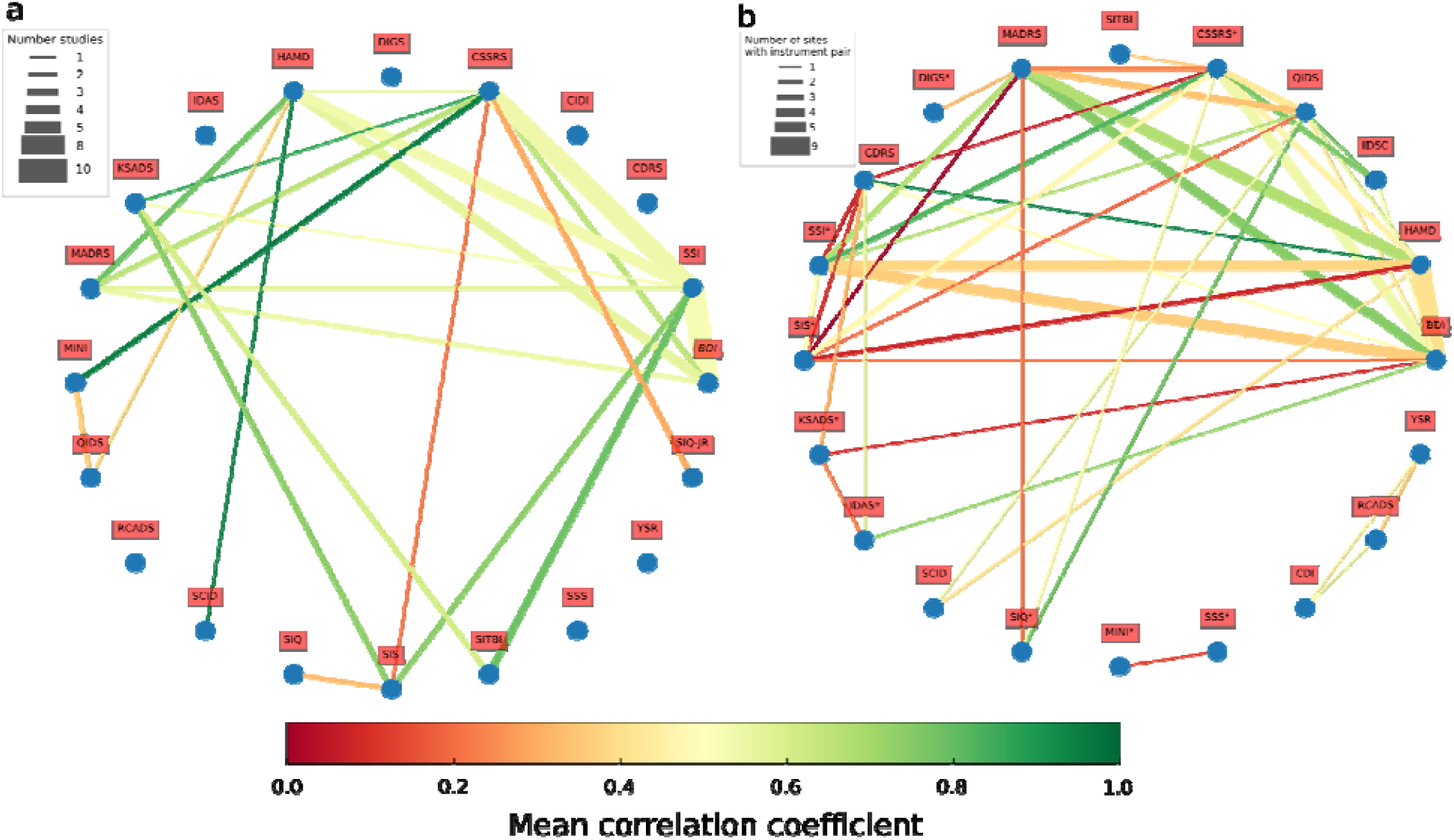
Overview of results. a) Literature review results. Reported instrument correlations are shown using an undirected graph. Nodes represent the instruments studied. Edges are coloured based on the average correlation across cohorts, edge width increases with the number of studies from which the correlations were extracted. b) ENIGMA correlation results. Each node represents one of the instruments included in the study. Edge color represents the sample-size weighted average correlation coefficient between two instruments. The thickness of the edge increases with the number of cohorts contributing to estimate the correlation. Generally speaking the thicker the edge the more confidence in the correlation estimate. c) Overview of the instruments used on the enigma analyses and the amount of overlap between them. BDI: Beck Depression Inventory suicidal ideation item; SSI: Scale for Suicidal Ideation; CDRS: Children’s Depression Rating Scale suicidal ideation item; CIDI: CIDI items on suicidal ideation and behaviour; C-SSRS: Columbia Suicide Severity Rating Scale; DIGS: Diagnostic Interview for Genetics Studies items on suicidal ideation and behaviour; HAM-D: Hamilton Depression Rating Scale item on suicidal ideation; IDAS-II: Inventory of Depression and Anxiety Symptoms suicide subscale; QIDS: Quick Inventory of Depressive Symptomatology suicidal ideation item; K-SADS: Kiddie Schedule for Affective Disorders and Schizophrenia suicide items; MADRS: Montgomery-Asberg Depression Rating Scale suicidal ideation item; MINI: Mini International Neuropsychiatric Interview suicidality module; RCADS: Revised Children’s Anxiety and Depression Scale suicidal ideation item; SCID: Structured Clinical Interview for DSM Disorders suicide questions; SIQ: Suicidal Ideation Questionnaire; SIS: Beck’s Suicide Intent Scale; SITBI: Self-Injurous Thouhts and Behaviours Interview; SSS: Suicide Score Scale; YSR: Youth Self-Report suicide item; SIQ-JR: Suicidal Ideation Questionnaire-Junior.

### ENIGMA meta-analysis

#### Sample description and dimensionality reduction

The average age across cohorts was 39 years (SD=16.3). Cohorts had on average 40% male participants. The most commonly available instruments were the suicidal ideation items from the MADRS, HAM-D and BDI questionnaires. Other relatively common instruments included the C-SSRS, QIDS, SSI, and SCID. The majority of instruments were administered by a clinician or trained interviewer, but some self-reported measures were used (**Table 2**). For complex instruments dimensionality reduction was carried out by extracting common factor scores using factor analysis (see methods). Fit statistics of these models for each complex instrument within each cohort is summarized in **Supplementary Table 3**. These complex instruments typically measure more than one suicidal construct. For example, the SSI includes sections on protective factors. Thus, a single common factor might not represent the best model underlying the latent structure of these instruments, but it serves our purpose of dimensionality reduction while capturing the main underlying latent factor related to suicidality which these instruments assess.

**Table 2.**
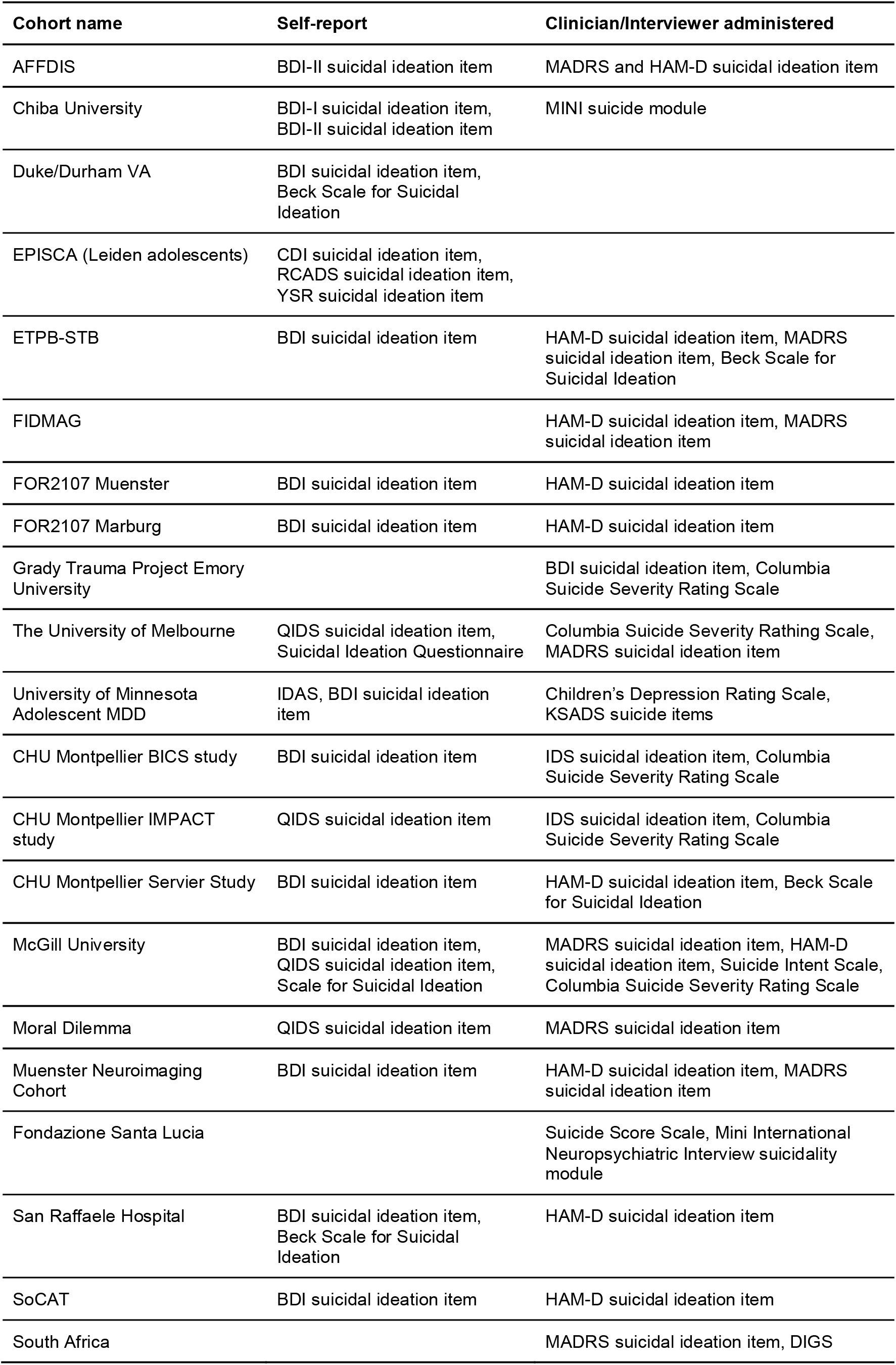
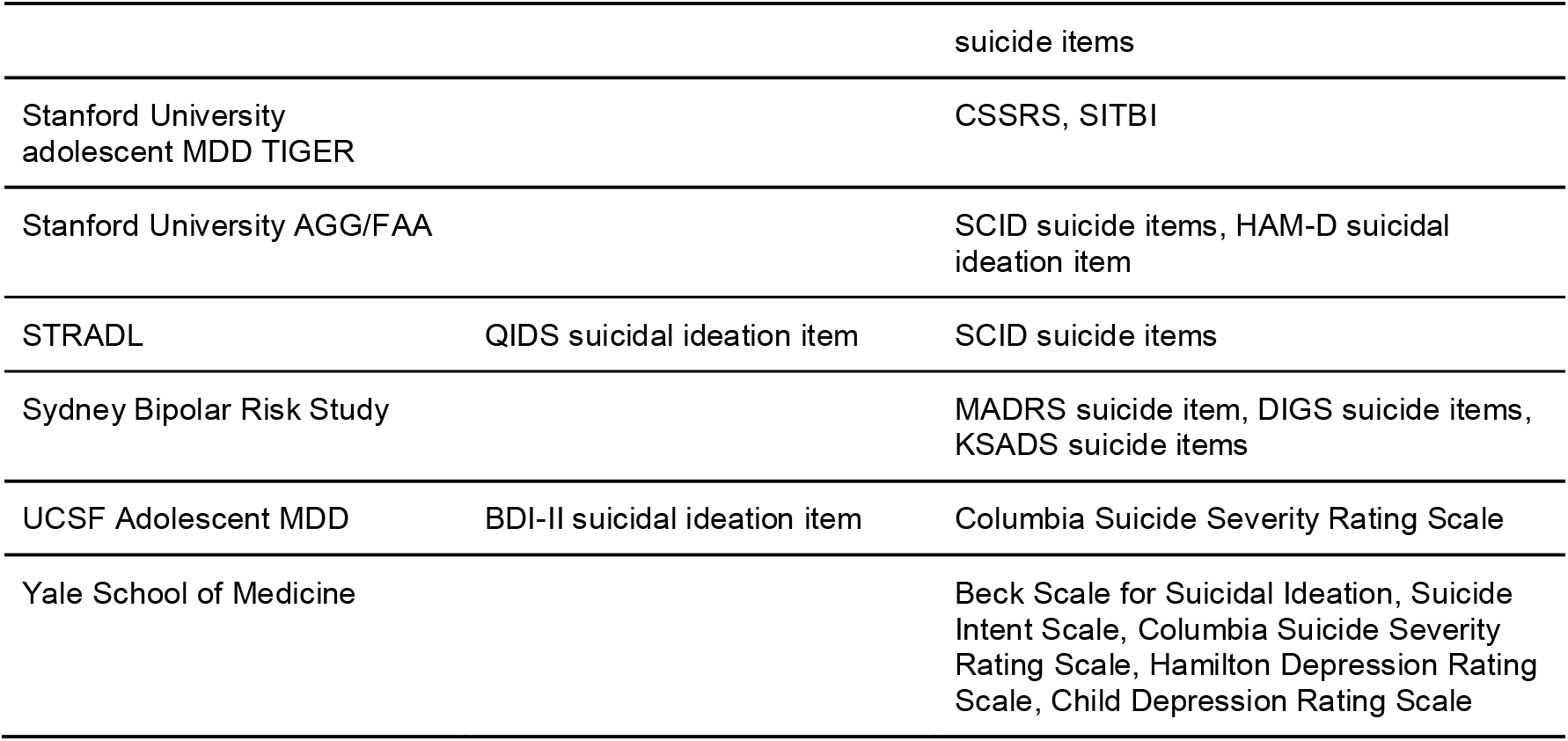
Instruments used to assess suicidal thoughts and/or behaviour by the different cohorts: self-report and clinician/interviewer administered measures are presented separately.

#### Correlation analyses

Results of our correlation analyses are summarised in **Figure 1b**. Full results are provided in **Supplementary Table S4**. As explained in the methods section, complex instruments were summarised using factor analysis for dimensionality reduction. From now on, when referring to the complex instruments listed in the methods, we are referring to the common factor score obtained by the dimensionality reduction approach. Overall moderate to high correlations (median r=0.44) were observed among all the studied instruments (including single-item and common-factor of complex instruments). Nonetheless, the common factor of the SIS showed poor correlations (median r∼0.20) with most of the instruments tested. This result is not unexpected; the SIS was applied by a single cohort (N=16) and assesses suicide intent during a suicide attempt, and not suicidal ideation or behaviour as the other instruments do (see discussion).

The instrument with the highest consistency (i.e., highest median weighted correlations with other instruments) was the IDS-Clinician rated measure (median r=0.76). However, few pairs of cohorts had data for this instrument. The C-SSRS and SSI instrument showed a very high concordance (r=0.83; N=191) with each other. In addition, there was a strong correlation between the HAM-D suicidal ideation item and the same item in the version of this questionnaire for children, the CDRS, but this was supported by a single cohort (r=0.94, N=20). The MADRS suicidal ideation item showed a high correlation with the HAM-D (r=0.67, N=1,087) and BDI (r=0.74, N=844) suicidal ideation items and with the SSI instrument common factor (r=0.67, N=119). The HAM-D and BDI suicide items showed only a moderate correlation (r=0.41, N=2,555) between them. Both of these measures were moderately correlated with the SSI (r=0.38, N=429 and r=0.36, N=350 respectively). Moderate to low correlations were observed for the group comprising child scales (YSR, RCADS and CDI), but these were only collected by one cohort. The MINI and SSS common factors showed a low correlation (r=0.12), which was also supported by a single cohort only (N=64).

#### Sensitivity analysis: recent versus lifetime STB

Cohorts applied different instruments assessing STB with different time frames. For example, the C-SSRS can be used to assess lifetime, time since last assessment, and recent (past 2 weeks) suicidal behaviour information, whereas other instruments might be worded around the past month, two weeks, week or even at the time of assessment. This is a potential source of heterogeneity for studies wishing to compare across these measures. For this reason, we repeated the analyses only focusing on measures applied to a *recent* (up to past month) time frame, and compared them to the results shown above. These analyses showed similar correlations overall. Notably, concordance between the C-SSRS and the HAM-D, as well as C-SSRS and BDI, were higher when focusing only on recent instruments (**Figure 2a-b** and **Supplementary Table S5**).

**Figure 2.**
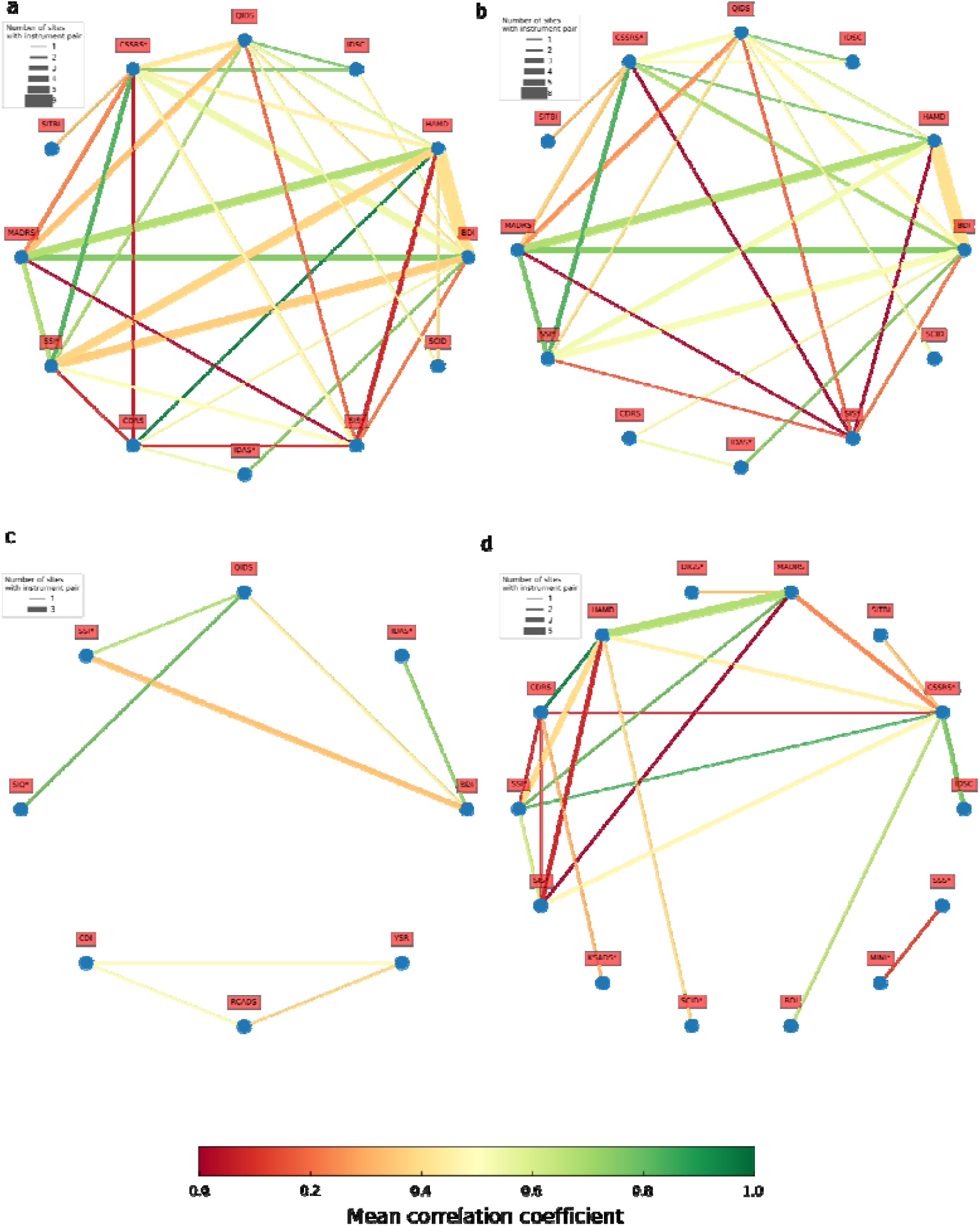
Sensitivity results. Undirected acyclic graph shows the results for the meta-analysis of correlations of suicide risk assessment instruments across ENIGMA cohorts for the complete results (a); using only measures assessing recent (up to past month) suicidal behaviour (b); instruments administered via self-report (c) or using clinician or interviewer based instruments (d; see Table 2). Each node represents one of the instruments included in the study. Each edge color represents the sample-size weighted average correlation coefficient between two instruments. The thickness of the edge increases with the number of cohorts contributing to estimate the correlation. Generally speaking the thicker the edge the more confidence in the correlation estimate. *For complex instruments dimensionality reduction was carried out by extracting common factor scores.

#### Comparing clinician and self-report scales

We gathered information on whether the distinct instruments were administered by a clinical interview or by self-report (**Table 2**), and the majority of instruments were administered by a clinician or trained interviewer. As a secondary sensitivity analysis, we repeated the analysis by including only either self-report or interviewer administered instruments (**Figure 2c-d**). Within interviewer-administered scales, high correlations (*r*>0.7) were observed between the SSI, C-SSRS, MADRS and QIDS instruments. A similar result was observed between HAM-D and MADRS (*r*=0.67). The SIS still showed a lower consistency with most other instruments (**Figure 2c**). For the self-report based instruments, less data was available. Among the self-reported instruments, the BDI and IDAS as well as the SSI and QIDS instruments showed a high concordance (**Figure 2d**). In general, higher correlations were seen for the interviewer-based measures (see **Supplementary Table S6** for the summary of the results). For interviewer-based measures, we were able to replicate the pattern from the main analyses: high correlations between single-item measures and measures assessing recent suicidal ideation (HAM-D, MADRS), strong correlations between detailed measures of STB (C-SSRS and SSI) and low correlations between the SIS and other measures. Nonetheless, we identified pairs of instruments such as the HAM-D and BDI whose low correlation in the main analysis might be explained by differences in administration (i.e., self-report vs interviewer).

## Discussion

Our study is a comprehensive assessment of how well different suicide risk assesment instruments relate to each other the extent to which they can be used interchangeably. Harmonization reduces heterogeneity and increases power for discovery analyses, but also enables the assessment of the generalisability of studies and opens up the opportunity to investigate other aspects such as interactions and individual variation analysis (van Harmelen et al., 2020). Identification of correlates of suicide risk may be improved by increasing sample sizes, and by pooling data across studies to detect small effect sizes, which may result from large variance in underlying mechanisms. Our study aimed to examine the concurrent validity of instruments commonly used to assess STBs. To this end, we compared individual level responses across questionnaires by pooling data from 27 cohorts belonging to the ENIGMA-MDD and ENIGMA-STB working groups. We compared our results to a systematic literature search across 180 studies.

A potential source of variance is the heterogeneity introduced by using different suicide risk assessment instruments that each measure slightly different underlying phenomena. Both the results of our analysis and our literature search identified moderate to high correlations between the most commonly used instruments to assess suicidal ideation including the BDI, SSI, HAM-D and MADRS, and between complex instruments (such as the C-SSRS and SSI). These findings are consistent with another study which showed strong correlations between the SSI, BDI and HAM-D (Desseilles et al. 2012). Nonetheless, our results were consistently more conservative than the literature (e.g., showing a lower degree of correlation). This could be explained in several ways. First, this might be evidence for publication bias whereby only positive and expected associations are published. Finally, heterogeneity arising from the way these instruments are administered (i.e. time frames for suicidal behaviours and self-report vs interview based) might also affect the results. We performed sensitivity analyses testing for these factors. For example, we proposed the low correlations between BDI and HAM-D may be explained by the fact that they are typically administered via self-report and a clinician interview respectively. There are studies reporting that the discrepancy between subjective and objective depression severity predicts differential treatment response in bipolar disorder (Suzuki et al., 2016). It is unclear whether a similar phenomenon underlies our results, given that the discrepancy is seen across cohorts rather than in individuals. Another example includes the correlation between the BDI and CSSRS common factor, which became higher when focusing on the interview based cohort only. Nonetheless, the heterogeneity explanation does not always hold true, for example, the MADRS (also interview-based) correlated well with the BDI.

Our unique methodology was chosen to integrate slightly different versions of instruments (i.e., self-reported vs. interview administered). By extracting common factor scores for the complex instruments, we were able to obtain a measure of the underlying suicidal liability being measured by the instruments while reducing the need to adjust for slightly different wording between versions. This approach might be more conservative than those used in previous studies as it is focused on measuring suicide liability rather than a specific construct such as attempt or ideation. As such, instruments that mainly focused on a specific aspect of STB such as suicidal ideation (for example the SIQ) are expected to show a lower correlation with more broad instruments that assess a range of relevant behaviors, such as the C-SSRS.

It is worth noting that suicidal constructs may share a common liability, but they may also have partially independent aetiologies. The Suicide Intent Scale (SIS) was the only measure with overall low measures of consistency in both our analyses and the literature search. However, caution is warranted in interpreting these findings as they were based on two cohorts only, which collected data on the SIS. Although our literature search identified a low concordance between the SIS and both the SIQ and the C-SSRS, a high concordance was reported between SIS and the K-SADS. These results are likely explained by the fact that the SIS is a questionnaire mainly focused on assessing intent of a past suicide attempt. This is further complicated by the intricate relationship between suicidal ideation, attempt, and actual suicide intent. Participants might engage in a suicide attempt with a relatively low intent to die. In fact, previous studies have identified that combining the SIS with other scales increases sensitivity and specificity for predicting suicide deaths (Stefansson et al., 2015).

Our study represents a comprehensive approach to assess the concordance and reliability of commonly used suicide risk assessment instruments. Nonetheless, some limitations need to be considered when interpreting our findings. Our literature search was as exhaustive and systematic as was practicable; however, we cannot rule out the possibility that some relevant studies were excluded because they are not indexed in the databases we searched. Language and cultural differences between cohorts might also affect whether two instruments are concordant. Our study comprised predominantly English speaking participants, but some cohorts included French, Dutch and German speaking participants. Limited research is available on whether language affects reporting of psychiatric symptomatology (Erkoreka et al., 2020). Participants might have undergone evaluation at different points in time in relation to the timing of suicidal behaviors or thoughts, and recall bias could lower the concordance between instruments. In fact, our sensitivity analysis showed that the time frame of the scale used can affect how different measures compare with each other. When focusing on complex instruments, we performed dimensionality reduction using factor scores derived from full-information maximum likelihood. Our approach was based on the fact that some instruments have multiple versions. Thus, our approach is not an exploratory or confirmatory factor analysis of the complex instruments used here. Performing such a study is outside the scope of this manuscript as it would require complete harmonisation of the questionnaires across cohorts and to focus solely on the complex instruments.

While our sensitivity analysis only distinguished between lifetime and recent time frames, it is possible that these effects exist even within recent time frames such as instruments assessing current vs. past two week behaviours. These limitations might explain the lower concordance identified by our analysis compared to the literature synthesis, which studied a compendium of smaller albeit less heterogeneous studies. Finally, while this was a large study, many sites had a distinct combination of measures collected (max N∼ 2,500); therefore, we did not have the power to perform additional sensitivity analyses in adults or adolescents only. Future studies should focus on addressing this, as there are clear factors associated with suicidality (e.g., mood reactivity) that are more prevalent during adolescence (Armey et al., 2015). We were also unable to stratify our analyses by type of psychiatric diagnosis, while prior work found that the correlation between self-report and clinician-reported suicidal thoughts may differ across disorders (Gao et al., 2015; Kaplan et al., 1994; Perugi et al., 2019).

Overall, our results suggest that the most commonly used instruments show a moderate to high concordance. Use of different measures of suicidality, might increase heterogeneity depending on the distinct dimensions and constructs assessed by each instrument. Our study could enable the implementation of composite scores by weighing more concordant measures heavily and penalising less concordant measures. In the absence of such an approach, large-scale collaborations could focus on strictly defined suicide constructs such as suicidal ideation, attempt, and intent that are preferentially defined using the most common instruments such as - amongst others - the HAM-D, MADRS, SSI, C-SSRS. In the absence of a common instrument, or when using an instrument that we found to have low concordance, sensitivity analyses could be performed to assess whether significant results are robust, or at least consistent, after excluding cohorts using the least common instruments. Future studies that plan to collect data on suicidal thoughts and behaviours would benefit from including one or more of the instruments that showed strong correlations with instruments such as the MADRS, SSI, and C-SSRS.

## Data Availability

NA

## Acknowledgements

MER received support from the Australian National Health and Medical Research Council (NHMRC) Centre for Research Excellence on Suicide Prevention (CRESP) [GNT1042580]. LS, LvV, LC, HPB, ALvH, SB and MD were supported by the MQ Brighter Futures Award MQBFC/2. LvV, LS and NJ were supported by the National Institute of Mental Health of the National Institutes of Health under Award Number R01MH117601. LS is supported by a NHMRC Career Development Fellowship (1140764). HPB was additionally supported by: R61MH111929RC1MH088366, R01MH070902, R01MH069747, American Foundation for Suicide Prevention, International Bipolar Foundation, Brain and Behavior Research Foundation, For the Love of Travis Foundation and Women’s Health Research at Yale. LC was additionally supported by Interdisziplinäres Zentrum für Klinische Forschung, UKJ. PMT was supported in part by NIH grant R01 MH116147. FJ would like to thank dr. S. Richard-Devantoy for his assistance during data collection. This work was supported by the CIBERSAM and the Catalonian Government (2014-SGR-1573 and 2017-SGR-1271 to FIDMAG). PF-C is funded by the Instituto de Salud Carlos III, co-funded by European Union (ERDF/ESF, “Investing in your future”): Sara Borrell contract (CD19/00149). The researchers based in Milan were supported by an Italian Ministry of Health grant RF-2011-02349921. ALvH was funded through the Leiden University Social Safety and Resilience Programme. The NIMH-ETPB team was supported by the Intramural Research Program at the National Institute of Mental Health, National Institutes of Health (IRP-NIMH-NIH; ZIAMH002927). MS is supported by the Phyllis and Jerome Lyle Rappaport Foundation, Ad Astra Chandaria Foundation, BIAL Foundation, Brain and Behavior Research Foundation, Anonymous donors, and the Center for Depression, Anxiety, and Stress Research at McLean Hospital. The team based at the University of Minnesota was supported by the National Institute of Mental Health (K23MH090421), the National Alliance for Research on Schizophrenia and Depression, the University of Minnesota Graduate School, the Minnesota Medical Foundation, and the Biotechnology Research Center (P41 RR008079 to the Center for Magnetic Resonance Research), University of Minnesota, and the Deborah E. Powell Center for Women’s Health Seed Grant. IHG was supported by the National Institute of Mental Health grant R37MH101495. EV thanks the support of the Spanish Ministry of Science and Innovation (PI15/00283, PI18/00805) integrated into the Plan Nacional de I+D+I and co-financed by the ISCIII-Subdirección General de Evaluación and the Fondo Europeo de Desarrollo Regional (FEDER); the Instituto de Salud Carlos III; the CIBER of Mental Health (CIBERSAM); the Secretaria d’Universitats i Recerca del Departament d’Economia i Coneixement (2017 SGR 1365), the CERCA Programme, and the Departament de Salut de la Generalitat de Catalunya for the PERIS grant SLT006/17/00357. NV thanks the support of a BITRECS project that has received funding from the European Union’s Horizon 2020 research and innovation programme under the Marie Skłodowska-Curie grant agreement No 754550 and from “La Caixa” Foundation (ID 100010434), under the agreement LCF/PR/GN18/50310006. For the team in Nimes, CINES grants access to HPC facilities (A0100311413). The team based in Rome was supported by an Italian Ministry of Health grant RC17-18-19-20-21/A. Support for the TIGER study includes the Klingenstein Third Generation Foundation, the National Institute of Mental Health (K01MH117442), the Stanford Maternal Child Health Research Institute, and the Stanford Center for Cognitive and Neurobiological Imaging. TCH receives partial support from the Ray and Dagmar Dolby Family Fund. The UCSF site was supported by the National Center for Complementary and Integrative Health (NCCIH) R21AT009173 and R61AT009864 to TTY; by the National Center for Advancing Translational Sciences (CTSI), National Institutes of Health, through UCSF-CTSI UL1TR001872 to TTY; by the American Foundation for Suicide Prevention (AFSP) SRG-1-141-18 to TTY; by UCSF Research Evaluation and Allocation Committee (REAC) and J. Jacobson Fund to TTY; by the National Institute of Mental Health (NIMH) R01MH085734 and the Brain and Behavior Research Foundation (formerly NARSAD) to TTY. The team at Duke University/Durham VA medical center were supported by the VA Mid-Atlantic Mental Illness Research Education and Clinical Center (MIRECC). The Moral Dilemma study was supported by the Brain & Behavior Research Foundation, and by the National Health and Medical Research Council (ID 1125504 to SLW). The FOR2107 Marburg site was supported by the German Research Foundation (DFG, grant FOR2107 KI588/14-1 and FOR2107 KI588/14-2 to Tilo Kircher, Marburg). The FOR2107 Muenster site was supported by the German Research Foundation (DFG, grant FOR2107 DA1151/5-1 and DA1151/5-2 to UD; SFB-TRR58, Projects C09 and Z02 to UD) and the Interdisciplinary Center for Clinical Research (IZKF) of the medical faculty of Münster (grant Dan3/012/17 to UD). CGD and BJH were supported by the National Health and Medical Research Council of Australia (NHMRC) Project Grants (1064643 and 1024570). NF was supported by the National Institute of Mental Health (MH111671) and National Center for Complementary and Integrative Health (R01AT011267). TJ was supported by the National Institutes Health Project Grants (MH098212) to TJ. JR and LF thank the support of the Spanish Ministry of Science and Innovation (PI19/00394 and CPII19/00009) integrated into the Plan Nacional de I+D+I and co-financed by the ISCIII-Subdirección General de Evaluación and the Fondo Europeo de Desarrollo Regional (FEDER); the Instituto de Salud Carlos III; the CIBER of Mental Health (CIBERSAM). YH, AN, ES were supported by AMED Brain/MINDS Beyond program Grant No. 20dm0307002, YH was supported by JSPS KAKENHI Grants No. 19K03309.

