## Supplements for "Concurrent validity and reliability of suicide risk assessment instruments: A meta analysis of 20 instruments across 27 international cohorts"

**Supplementary Table 1. Reliability of items, scales, questionnaires and interviews assessing suicidal thoughts and behaviours**

| Measure | Description | Focus | Inter-rater reliability | Internal consistency | Test-retest reliability |
| --- | --- | --- | --- | --- | --- |
| Beck Depression Inventory  suicide item;  [1][2] | Item on suicidal ideation in self-report depression severity questionnaire  (scored 0-3) | Suicidal ideation in the past two weeks for BDI-II, past week for BDI-I | NA | NA | 40% of adolescents who scored 2 or 3 on the BDI suicide ideation item still reported significant suicidal ideation (a score of 2 or 3) when retested 4- to 6-weeks later [3] In a sample with clinically suicidal young adults, the 6-month test–retest correlation for the BDI suicide item was .35 [4] |
| Scale for Suicidal Ideation (SSI) (interviewer-administered) & Beck Scale for Suicidal Ideation (BSSI) (self-report);  [5][6] | 19 item clinician-rated semi-structured interview (SSI) or self-report questionnaire (BSSI) about intensity of (active or passive) ideation and intent to end life by suicide (all items are scored 0-2). An additional 2 items are not scored | Suicidal ideation in the past week | Interrater reliability for the SSI ranges between .83-.89 [7, 8]. Intra-class correlation for the SSI was .97 [9] | Cronbach's alpha: .84 - .97 [5, 10][11][6]  Cronbach’s alpha varied between .75 and .96 for translated versions [12, 13] | Three month test-retest reliability was .74 [14]. One-week test-retest reliability was r = .88 [11] and r = .54 [15] |
| Children's Depression Rating Scale Revised (CDRS-R) suicide item [16] | Clinician-rated interview to assess depression severity in children, based on the Hamilton Depression Rating Scale and includes 17 items. Includes one item on suicidal ideation and attempt (scored 1-7) | Suicidal ideation and suicidal behaviour in the past week | Acceptable interrater reliability was observed for the suicide index (K = .74) [17]. Interrater reliability for the suicide ideation scale was kappa=.65 in both clinical and non-clinical samples [16] | NA | NA |
| Composite International Diagnostic Interview (CIDI) suicide questions [18] | Suicidal ideation, plans, and behaviour are assessed in the major depressive disorder module of the CIDI | Lifetime suicidal ideation and behaviour | NA | NA | NA |
| Columbia-Suicide Severity Rating Scale (C-SSRS) [19] | Semi-structured interview to assess recent and lifetime suicidal ideation and suicidal behaviour, including severity and intensity of ideation and lethality of attempt. The intensity and severity scales are scored on a five-point ordinal scale | Recent and lifetime suicidal ideation, suicidal behaviour, including lethality of attempts | Interrater reliability: weighted kappa of .92 and .88 for recent and lifetime most severe ideation respectively in a Turkish sample [20]. Kappa was .75 for lifetime attempt, 1 for lifetime ideation, .84 for suicidal ideation in the past month and .52 for attempt in the past month in a sample of inpatients [21]. The weighted kappa for suicidal behaviour ranged from .70-.90 [22]. ICC=.09 for suicidal ideation, and 100% agreement for suicidal behaviour in adolescent sample [23]; k=.88 for distinguishing actual, aborted, preparatory acts and other acts in adolescent sample [24] | Cronbach’s alpha: In the original study by Posner et al., 2011 Cronbach’s alpha was between .73 and .93 [19].  Alpha was .87 for ideation subscale, .73 suicide intensity scale,.89 for severity subscale and .91 for behaviour subscale in Spanish adolescents [25]. In a Spanish clinical sample, Cronbach’s alpha was .53 for the intensity subscale [26]. Cronbach’s alpha was .89 and .91 for a recent and lifetime C-SSRS score in a Turkish sample [20], .95 in an inpatient sample [27]. Alpha was .53 for lifetime scores and .50 for past month in a sample of inpatients [21] and .64 for the five initial questions in a sample of adult psychiatric patients [22]. | NA |
| Diagnostic Interview for Genetic Studies (DIGS) suicide items [28] | Diagnostic interview which includes items about lifetime suicidal ideation and suicidal behaviour | Lifetime suicidal ideation and behaviour | The inter-rater agreement on suicidal behaviour using a Portuguese translation of the DIGS was 96.4% [29] | NA | NA |
| Hamilton Depression Rating Scale (HDRS)  suicide item [30] | Clinician-rated scale with one item on suicidal ideation and attempt, which is scored from 0 (no ideation) to 4 (suicide attempt) | Suicidal ideation and behaviour in the past week | Inter-rater reliability: .95 [31] and .90 [32] | NA | Test-retest reliability in a three day period is r=.64 [33] |
| Inventory of Depression and Anxiety Symptoms (IDAS-II) - Suicide Subscale [34] | Questionnaire to assess depressive and anxiety symptoms that includes 6 items on suicidal ideation and behaviour  (scored 1-5) | Suicidal ideation and behaviour in the past two weeks | NA | Cronbach’s alpha for the suicidality scale range between .79 and .86 in young adults, high school students, college student and patients [34] In a large community sample, Cronbach’s alpha for the suicide subscale was .77 [35] | Test-retest reliability in patients was .77 for the IDAS-I suicidality scale for a 1-week time period [36] |
| Inventory of Depressive Symptomatology (IDS) & Quick Inventory of Depressive Symptomatology (QIDS) suicide item  [37] | Depressive symptom severity questionnaire that includes one item on suicidal ideation and/or behaviour (scored 0-3) | Suicidal ideation and behaviour in the past week | NA | NA | NA |
| Kiddie Schedule for Affective Disorders and Schizophrenia (K-SADS) suicide items [38] | Semi-structured diagnostic interview with five questions in the MDD module around ideation, suicidal behaviour and non-suicidal self-injury (scored 0-3) | Lifetime suicidal ideation and behaviour, non-suicidal self-injury | Interrater reliability: k=.9 for ideation, .83 for attempt and .71 for NSSI [39] | NA | NA |
| Montgomery–Åsberg Depression Rating Scale (MADRS) suicide item [40] | Clinician-administered depressive symptom severity assessment, with one item on suicidal ideation (scored 0-6) | Suicidal ideation in the past week | Interrater reliability: spearman correlation=.63 [41] ICC ranged between 0.95 and 0.99 for three and two raters respectively on a Japanese version of the MADRS [42] ICCs were lower than 0.60 in three different samples [43] | NA | NA |
| Mini International Neuropsychiatric Interview (MINI) suicidality module [44] | Diagnostic interview which includes 9 items around suicidal ideation, self-harm and suicidal behaviour (scored yes/no). | Suicidal ideation and behaviour, non-suicidal self-injury in the past month & lifetime | NA | NA | NA |
| Revised Children's Anxiety and Depression Scale (RCADS) suicide item [45] | Questionnaire to assess severity of depressive and anxiety symptoms in children, which includes one item on frequency of thoughts about death (scored on a four-point likert scale) | Lifetime suicidal ideation | NA | NA | NA |
| Structured Clinical Interview for DSM Disorders (SCID) Mood Disorder Module Suicide Questions [46] | Structured clinical interview that includes items on suicidal ideation, suicide plans and suicidal behaviour during depressive episodes (scored 1-3) | Lifetime suicidal ideation and behaviour | NA | NA | NA |
| Suicidal Ideation Questionnaire (SIQ) [47] | 30-item self-report questionnaire to assess suicidal ideation in adolescents  (grades 10-12)  (scored 0-6 per item) | Lifetime suicidal ideation | NA | Cronbach's alpha: .97 in the original clinical adolescent sample [47]. Cronbach’s alpha was .975 for a Chinese version in adolescents [48], .95 for Kuwaiti students and .96 for American students [49], .973 in Chinese high school students [50], .95 in French adolescents [51] and .97 in a clinical adolescent sample [52] | Test retest correlation was .91 for a two-week period in French adolescents [51]. In a large sample of high school students, the SIQ had a test-retest reliability, over an interval of approximately 4 weeks, of .72 [53] |
| Beck's Suicide Intent Scale (SIS) [54] | Clinican-rated Interview with 15 items to assess the seriousness of previous suicide attempts (scored 0-2). Questions detail the preparation, lethality, expectations and planning. An additional 5 items are not scored | Suicide intent during most recent attempt and attempt with highest lethality | Interrater reliability: .81-.95 [54, 55]. | Cronbach’s alpha: .95 [54], and .651 in Chinese samples [56]. Alpha was .85 in a sample of adolescent suicide attempters [57]. Alpha was .84 in a clinical sample [58] and in two samples of adolescents with recent attempts (α=.74 and .79) [59, 60] | NA |
| Self-Injurious Thoughts and Behaviours Interview  (SITBI) [39] | Structured interview that includes 169 items on suicidal ideation, plans, suicidal behaviour and non-suicidal self-injury | Suicidal ideation and behaviour, non-suicidal self-injury during lifetime, past year and past month | Interrater reliability: k= .99 [39], perfect for ideation, plans, gestures, attempts, NSSI in the previous year and month in a Spanish population [61], A German version showed good interrater agreement for NSSI (.77) and perfect agreement for suicidal behaviour [62] | NA | Test retest reliability: k=.7-1 for suicidal ideation, plan, attempt and NSSI over six months, but poor for suicidal gestures (K=.25) [39]. |
| Suicide Score Scale (SSS) [63] | 20-item self-report questionnaire to assess suicidal ideation and behaviour | Suicidal ideation and behaviour during past year and lifetime | NA | Cronbach's alpha: .75 for part I (ideation and attempt in last 12 months) and .80 for part II (lifetime ideation and attempt) in undergraduate students [63], and .87 in a different student sample [64]. | NA |
| Youth Self-report suicide item [65] | Self-report questionnaire to assess internalizing and externalizing behaviour, which includes an item on suicidal behaviour (deliberate self-harm or suicidal behaviour;  scored 0-2) | Suicidal ideation and behaviour, non-suicidal self-injury in the past six months | NA | NA | NA |
| Suicidal Ideation Questionnaire JR (SIQ-JR) [47] | 15-item self-report version of the SIQ for grade 7-9  (scored 0-6) | Lifetime suicidal ideation |  | Cronbach's alpha: .93, .94, .94 for seventh, eighth, and ninth graders [53] .951 for Chinese version in adolescents [48] and ranged between .93-.95 for American adolescents [66], .92 in a sample of inpatient adolescents [67], and .96 in a sample of American Indian adolescents [68] and .91 in inner-city adolescents [69] | Three-week test-retest reliability was r=.89 in inner-city children and adolescents [69] |

**Supplementary Table 2. Concurrent and predictive validity of items, scales, questionnaires and interviews assessing suicidal thoughts and behaviours**

| **Measure** | **Concurrent validity** | **Predictive validity** |
| --- | --- | --- |
| Beck Depression Inventory item 9 | Correlation between BDI suicide item and BSSI was r=.56-.58 [15]  Correlation was .48 with the first five items of the SSI in hospitalized suicide attempters [70]  Correlation was .68 with the total BSSI score in university students  [14] and .69 in adolescents [71]  Correlation with BSI score was .41 in an inpatient sample and .69 in an outpatient sample [5]  Correlation was .58 with the SSI total score and .53 with the Hamilton Depression Rating Scale item on suicidal ideation [72].  Agreement on presence of ideation between the BDI (score 2 or higher) and Hamilton Depression Rating Scale suicidal ideation item (score 3 or higher) was kappa=.64 in primary care patients, kappa=.52 in outpatient and kappa=.39 in inpatients  [73]  The BDI suicide item was correlated with the MADRS and HDRS suicide items (r=.65 and .69) [74]  Correlations between the BDI suicide item and SSI score were r=.69 for outpatients and r=.58 for inpatients [6] | Participants who scored higher than 2 on the BDI suicidal ideation item, were 6.9 times more likely to commit suicide than those that scored lower [75]  BDI item 9 showed good positive predictive value (.53) for suicidal behaviour in six months following the interview, but low sensitivity (.33) [72]  BDI score predicted death by suicide and repeat suicide attempts in an outpatient sample with follow-up between 18 months and 20 years [76].  In a community sample of adolescents, the BDI suicidal ideation item was found to be predictive of both future suicide attempts (OR=6.9) and future depressive episodes (OR=2.1) [77]  A non-zero score on the BDI suicidal ideation item predicted death by suicide in an impatient sample followed for 30 years (OR=2.41) [78] |
| Scale for Suicidal Ideation (SSI) (interviewer-administered) & Beck Scale for Suicidal Ideation (BSI/BSSI) (self-report) | SSI total scores correlated moderately with BDI suicide item score (r=.58) and Hamilton Depression Rating Scale suicidal ideation item (r=.67) [72].  Agreement on presence of ideation between the SSI sum score (six or higher) and the BDI item 9 (score 2 or higher) was higher in a primary care sample (kappa=.45), then in outpatients (kappa=.29) and inpatients (kappa=.32), while agreement on the presence of ideation between the SSI sum score (six or higher) and HDRS item 3 (score 2 or higher) was highest in outpatients (kappa=.50), followed by primary care (kappa=.49) and inpatients (kappa=.42) [73]  Correlation between BSSI scores and the SSI scores was r=.90, and correlation between the BDI suicide item and SSI were .69 for outpatients and r=.58 for inpatients [6]  Agreement on presence of ideation between SITBI and BSSI was K=.59 [39]  The BSSI scores were correlated with the BDI suicide item (r=.69) [71] | Outpatients with a total score higher than 2 on the SSI, were 7 times more likely to commit suicide than those that scored lower [79].  In a sample of more than 3000 adult outpatients, those who had a total score higher than 2 on the SSI had 5.42 times higher odds of dying by suicide than under [80].  In psychiatric outpatients, those that scored 16 or higher on the SSI during their worst ever suicidal ideation, were almost 14 times more likely to die by suicide than those that scored below 16 [80] |
| Children's Depression Rating Scale (CDRS) suicide item | NA | A suicide index composed of the sum of responses to five measures of suicidality (including the CDRS suicidal ideation item) was moderately associated with suicidal ideation one year later (r=.39) [81]  Score on the CDRS-R suicidal ideation item was not related to suicidal behaviour during 36-week treatment in adolescents [82] |
| Composite International Diagnostic Interview (CIDI) suicide questions | NA | NA |
| Columbia-Suicide Severity Rating Scale (C-SSRS) | C-SSRS severity scores were correlated with the HDRS suicide item (r=.56) and with Beck Suicide Intent Scale scores in those with suicide attempt (r=.22) [26]  C-SSRS severity scores and intensity scores were also correlated with SSI worst-point scores (r=.52 and r=.56 respectively) [19]  The C-SSRS severity scores were correlated with the MADRS suicide item (r=.63) and the BDI suicide item (r=.80), while the intensity scores were also correlated with the MADRS item (r=.69) and BDI item (r=.51) [19]  There were modest correlations between the C-SSRS severity scale and SIQ-JR total score (r=.36) and between C-SSRS intensity subscale and SIQ-JR total score (r=.23) [19]  The correlation between C-SSRS severity and SSI total scores were moderate (r=.69), the correlation between C-SSRS intensity subscale scores and SSI total scores was modest (r=.34) [19]  C-SSRS scores were associated with SSI scores (r=.71) and KSADS ideation item (r=.87) in Spanish adolescents [25]  The total C-SSRS score was correlated with the BSSI score (r=.477) in an inpatient sample [27]  In a sample of veterans, the severity and ideation subscale were moderately correlated with the SSI total score (r=.50 and .52) [83]  Agreement between the MINI and C-SSRS for lifetime suicidal behaviour (k=.97) [84] | C-SSRS intensity scale score predicted suicide attempt at one-year follow-up in adolescents (OR=1.09). Duration of ideation was a predictor of suicide attempt at follow-up (OR=1.80) [85]  Baseline C-SSRS severity scores based on worst-point lifetime ideation were significantly predictors of suicide attempt in the following six months (OR=1.45) [19]  In adolescents, suicide attempt reported at baseline predicted suicidal behaviour at follow-up three months later (OR=4.03), but severity and intensity of suicidal ideation at baseline did not predict future attempt over and above baseline attempt [86]  The C-SSRS total score was associated with suicidal behaviour in the six months following hospitalization (r between .239 and .289) [27].  C-SSRS total score and intensity score predicted a suicide attempt in the following six months (OR=1.08 and 1.1 respectively) in an adult sample with psychiatric illness [22]  In a veteran sample, baseline intensity subscale and severity subscale scores predicted attempt at 6 months (OR between 1.19-3.23) [83]  Severity of ideation and history of suicide attempt predicted suicide attempt in the next 18 months in young people (OR=1.51 and 4.80 respectively), in ideators intensity and frequency of ideation predicted suicide attempt (OR=1.15 and 1.54 respectively) [87]  Suicidal ideation and suicidal behaviour reported on the electronic version of the C-SSRS predicted suicide attempt in psychiatric and non-psychiatric patients [88].  Current and worst-ever severity of ideation predicted future suicidal behaviour 12 months after visiting an emergency department for suicide risk [89] |
| Diagnostic Interview for Genetic Studies (DIGS) suicide items | NA | NA |
| Hamilton Depression Rating Scale (HDRS) (interviewer-administered) suicide item | The HDRS suicide item was correlated with the first five items of the SSI (r=.40) [70]  HDRS item 3 was correlated with C-SSRS severity score in a Spanish sample (r=.56) [26]  HDRS and MADRS suicide items were strongly correlated in a ketamine trial for treatment-resistant major depressive disorder (r=.86) [74]  HDRS suicide item scores were moderately associated with BDI suicide item scores (r=.53), and total SSI scores (r=.67) [72]  When using a cut-off, agreement between the SSI (6 or higher) and HDRS suicide item (2 or higher) was stronger (kappa=.70) than agreement between SSI (6 or higher) and BDI (2 or higher) (kappa=.15) [72]  There was a strong concordance between SCID suicide items and HDRS suicide item in older participants (r=.94) [90]  The agreement between QIDS and HDRS was 76% (kappa=.40) [91] | Participants who score 2 or higher were 4.9 times more likely to die by suicide than those that scored lower [75] |
| Inventory of Depression and Anxiety Symptoms (IDAS) - Suicide Subscale | The IDAS suicidality scale was significantly correlated with the suicidality items from the Interview for Mood and Anxiety Symptoms (r=.62) [36] | NA |
| Kiddie Schedule for Affective Disorders and Schizophrenia (K-SADS) suicide items | When the KSADS suicide items were used to categorize patients, the Beck SSI scores significantly differed between all five classes [92]  The KSADS suicide items correlated with the SSI (r=.52) and SIS scores (r=.76) [93]  Good agreement between SITBI and KSADS on presence of suicide attempt (K=.65), and NSSI (K=.74), but lower agreement on ideation (K=.48) [39] | Higher suicidal ideation at baseline on the K-SADS in adolescents was associated with suicide attempt in young adulthood in girls (OR=8.81), but not boys [94] |
| Montgomery–Åsberg Depression Rating Scale (MADRS) suicide item | The MADRS suicide item score correlated with the first five items of the SSI r=.62, BDI suicide item r=.45, HDRS suicide item r=.71 and SSI total score r=.53 [74] | NA |
| Mini International Neuropsychiatric Interview (MINI) suicidality module | Agreement between MINI suicide items and QIDS item was 83.5% (weighted kappa: .30) in MDD and 83.1% (weighted kappa: .43) in BD [95]  There was a significant correlation between C-SSRS scores and the suicidality score from the MINI-KID (Children and adolescent version of the MINI) (r=.94) in a Turkish adolescent sample [20] | The shorter 6-item version of the MINI suicide scale significantly predicted suicidal behaviour, but not NSSI 3 and 12 months after discharge (OR=3.1-8.2 for the different items) in psychiatric patients [96]  Suicidality subscale scores predicted suicide attempt at two-year follow-up in homeless people with mental illness (Log OR=1.08) [97] |
| Quick Inventory of Depressive Symptomatology (QIDS) suicide item | Agreement between MINI suicide items and QIDS item was 83.5% (weighted kappa: .30) in MDD and 83.1% (weighted kappa: .43) in BD [95]  The agreement between QIDS and HDRS suicide items was 76% (kappa=.40) [91] | NA |
| Revised Children's Anxiety and Depression Scale (RCADS) suicide item | NA | NA |
| Structured Clinical Interview for DSM Disorders (SCID) Mood Disorder Module Suicide Questions | Strong concordance between SCID suicide items and HDRS suicide item in older participants (r=.94) [90] | NA |
| Suicidal Ideation Questionnaire (SIQ) | The SIQ scores were correlated with the SIS scores in adolescents (r=.26) [57] | NA |
| Beck's Suicide Intent Scale (SIS) | SIS scores were positively correlated with C-SSRS severity scale scores in a Spanish outpatient sample (r=.22) [26]  Negative association with SIQ (r=-.39) [98]  Correlations between the SIS and SSI were r=.73 and r=.76 in prisoners and young adolescents respectively [93, 99]  SIS scores were correlated with scores from the KSADS in children and young adolescents (r=.72) [93]  SIS total scores correlated with the SIQ (r=.26) in adolescents [57] | SIS did not predict suicide 5-10 years later [100]  SIS did not predict suicide 1.5-4 years later [101]  SIS predicted suicide 12 years later in a Finnish sample (OR=1.2) [102]  SIS score above 19 predicted suicide at follow up (median 6 years later) in women only [103]  SIS did not predict suicidal behaviour in psychiatric patients in the following 10 years [104]  SIS scores predicted death by suicide at follow-up (mean follow-up 5.2 years later), but the positive predictive value of the SIS was low (4%) [105]  SIS scores distinguished those who died by suicide at follow-up (mean follow-up 9.5 years later) and those that did not (positive predictive value = 19%)[106] |
| Self-Injurious Thoughts and Behaviours Interview  (SITBI) | Good agreement between SITBI and KSADS on presence of suicide attempt (K=.65), and NSSI (K=.74), but lower agreement on ideation (K=.48) [39]  Agreement between the Spanish version of the SITBI and the Beck Scale for Suicidal Ideation was k=.99 for ideation, suicide plans and suicide attempt, but lower (k=.78) for suicidal gestures [61]  Agreement on presence of ideation between SITBI and BSSI was good (K=.59) [39] | NA |
| Suicide Score Scale (SSS) | Moderate negative correlation with the Reasons for Living Scale (r=-.32) and Zung Depression Scale (r=-.41) [63] | NA |
| Youth Self-report suicide item | NA | A suicide index composed of the sum of responses to five measures of suicidality (including the YSR and the CBCL suicidal ideation/behaviour items) was moderately associated with suicidal ideation one year later (r=.39) [81] |
| Suicide Ideation Questionnaire JR (SIQ-JR) | NA | SIQ-JR predicted suicidal ideation or behaviour 6 months after discharge (OR=1.84) and suicide attempt in the follow-up period in inpatient adolescents (OR=1.74) [66]  SIQ-JR was a significant predictor of attempt two months later in American Indian Adolescents [68]  In adolescent inpatients, higher SIQ-JR scores were predictive of suicidal thoughts and behaviour six months after hospitalization [107]  SIQ-JR scores predicted suicide attempt one year later in girls, but not in boys [67] |

| **Supplementary Table 3. Fit statistics for the common-factor models used for dimensionality reduction** | | | |
| --- | --- | --- | --- |
| **Cohort** | **Instrument** | **CFI** | **RMSEA** |
| **The University of Melbourne** | **CSSRS** | **0.639** | **0.139** |
| **The University of Melbourne** | **SIQ** | **0.787** | **0.112** |
| **San Rafaelle Hospital** | **SSI** | **0.589** | **0.109** |
| **ETPB-STB** | **SSI** | **2.521** | **0** |
| **Fondazione Santa Lucia** | **MINI** | **0.876** | **0.071** |
| **Fondazione Santa Lucia** | **SSS** | **0.558** | **0.108** |
| **UCSF Adolescent MDD** | **CSSRS** | **0.326** | **0.131** |
| **CHU Montpellier Servier Study** | **SSI** | **0.264** | **0.266** |
| **Duke/Durham VA** | **SSI** | **0.332** | **0.152** |
| **Grady Trauma Project Emory University** | **CSSRS** | **1.054** | **0** |
| **McGill University** | **CSSRS** | **0.671** | **0.178** |
| **McGill University** | **SSI** | **0.687** | **0.097** |
| **McGill University** | **SIS** | **0.433** | **0.159** |
| **CHU Montpellier BICS study** | **CSSRS** | **0.614** | **0.164** |
| **Sydney Bipolar Risk Study** | **DIGS** | **0.981** | **0** |
| **Sydney Bipolar Risk Study** | **KSADS** | **0.965** | **0** |
| **Stanford University adolescent MDD TIGER** | **CSSRS** | **0.446** | **0.198** |
| **Stanford University AGG/FAA** | **SCID** | **0.992** | **0.092** |
| **CHU Montpellier IMPACT study** | **CSSRS** | **0.448** | **0.311** |
| **University of Minnesota adolescent MDD** | **KSADS** | **0.965** | **0.091** |
| **University of Minnesota adolescent MDD** | **IDAS** | **0.974** | **0.161** |
| **Yale School of Medicine** | **CSSRS** | **0.513** | **0.193** |
| **Yale School of Medicine** | **SSI** | **0.863** | **0.07** |
| **Yale School of Medicine** | **SIS** | **0.657** | **0.074** |

**Supplementary Table 4. Correlations between different suicide measures**

|  | BDI | HAMD | IDSC | CSSRS* | MADRS | CDRS | KSADS* | IDAS* | QIDS | SSI* | SIS* | SCID | SIQ* | SITBI | DIGS* | MINI* | SSS* | CDI | RCADS | YSR |
| --- | --- | --- | --- | --- | --- | --- | --- | --- | --- | --- | --- | --- | --- | --- | --- | --- | --- | --- | --- | --- |
| HAMD | 0.41 (s.d.=0.17; Ncohorts=9) |  |  | 0.46 (s.d.=0.12; Ncohorts=2) | 0.67 (s.d.=0.16; Ncohorts=5) | 0.94 (s.d.=0.00; Ncohorts=1) |  |  | 0.56 (s.d.=0.00; Ncohorts=1) | 0.38 (s.d.=0.22; Ncohorts=5) | 0.08 (s.d.=0.19; Ncohorts=2) | 0.42 (s.d.=0.00; Ncohorts=1) |  |  |  |  |  |  |  |  |
| IDSC | 0.51 (s.d.=0.00; Ncohorts=1) |  |  | 0.76 (s.d.=0.20; Ncohorts=2) |  |  |  |  | 0.77 (s.d.=0.00; Ncohorts=1) |  |  |  |  |  |  |  |  |  |  |  |
| CSSRS* | 0.54 (s.d.=0.08; Ncohorts=4) | 0.46 (s.d.=0.12; Ncohorts=2) | 0.76 (s.d.=0.20; Ncohorts=2) |  | 0.24 (s.d.=0.20; Ncohorts=2) | 0.06 (s.d.=0.00; Ncohorts=1) |  |  | 0.43 (s.d.=0.11; Ncohorts=3) | 0.83 (s.d.=0.01; Ncohorts=2) | 0.47 (s.d.=0.05; Ncohorts=2) |  | 0.50 (s.d.=0.00; Ncohorts=1) | 0.35 (s.d.=0.00; Ncohorts=1) |  |  |  |  |  |  |
| MADRS | 0.75 (s.d.=0.02; Ncohorts=4) | 0.67 (s.d.=0.16; Ncohorts=5) |  | 0.24 (s.d.=0.20; Ncohorts=2) |  |  |  |  | 0.34 (s.d.=0.23; Ncohorts=3) | 0.67 (s.d.=0.10; Ncohorts=2) | -0.13 (s.d.=0.00; Ncohorts=1) |  | 0.22 (s.d.=0.00; Ncohorts=1) |  | 0.33 (s.d.=0.00; Ncohorts=1) |  |  |  |  |  |
| CDRS | 0.49 (s.d.=0.00; Ncohorts=1) | 0.94 (s.d.=0.00; Ncohorts=1) |  | 0.06 (s.d.=0.00; Ncohorts=1) |  |  | 0.32 (s.d.=0.00; Ncohorts=1) | 0.54 (s.d.=0.00; Ncohorts=1) |  | 0.08 (s.d.=0.00; Ncohorts=1) | 0.10 (s.d.=0.00; Ncohorts=1) |  |  |  |  |  |  |  |  |  |
| KSADS* | 0.08 (s.d.=0.00; Ncohorts=1) |  |  |  |  | 0.32 (s.d.=0.00; Ncohorts=1) |  | 0.24 (s.d.=0.00; Ncohorts=1) |  |  |  |  |  |  |  |  |  |  |  |  |
| IDAS* | 0.73 (s.d.=0.00; Ncohorts=1) |  |  |  |  | 0.54 (s.d.=0.00; Ncohorts=1) | 0.24 (s.d.=0.00; Ncohorts=1) |  |  |  |  |  |  |  |  |  |  |  |  |  |
| QIDS | 0.45 (s.d.=0.00; Ncohorts=1) | 0.56 (s.d.=0.00; Ncohorts=1) | 0.77 (s.d.=0.00; Ncohorts=1) | 0.43 (s.d.=0.11; Ncohorts=3) | 0.34 (s.d.=0.23; Ncohorts=3) |  |  |  |  | 0.68 (s.d.=0.00; Ncohorts=1) | 0.19 (s.d.=0.00; Ncohorts=1) | 0.52 (s.d.=0.00; Ncohorts=1) | 0.77 (s.d.=0.00; Ncohorts=1) |  |  |  |  |  |  |  |
| SSI* | 0.37 (s.d.=0.27; Ncohorts=5) | 0.38 (s.d.=0.22; Ncohorts=5) |  | 0.83 (s.d.=0.01; Ncohorts=2) | 0.67 (s.d.=0.10; Ncohorts=2) | 0.08 (s.d.=0.00; Ncohorts=1) |  |  | 0.68 (s.d.=0.00; Ncohorts=1) |  | 0.51 (s.d.=0.16; Ncohorts=2) |  |  |  |  |  |  |  |  |  |
| SIS* | 0.22 (s.d.=0.00; Ncohorts=1) | 0.08 (s.d.=0.19; Ncohorts=2) |  | 0.47 (s.d.=0.05; Ncohorts=2) | -0.13 (s.d.=0.00; Ncohorts=1) | 0.10 (s.d.=0.00; Ncohorts=1) |  |  | 0.19 (s.d.=0.00; Ncohorts=1) | 0.51 (s.d.=0.16; Ncohorts=2) |  |  |  |  |  |  |  |  |  |  |
| BDI |  | 0.41 (s.d.=0.17; Ncohorts=9) | 0.51 (s.d.=0.00; Ncohorts=1) | 0.54 (s.d.=0.08; Ncohorts=4) | 0.75 (s.d.=0.02; Ncohorts=4) | 0.49 (s.d.=0.00; Ncohorts=1) | 0.08 (s.d.=0.00; Ncohorts=1) | 0.73 (s.d.=0.00; Ncohorts=1) | 0.45 (s.d.=0.00; Ncohorts=1) | 0.37 (s.d.=0.27; Ncohorts=5) | 0.22 (s.d.=0.00; Ncohorts=1) |  |  |  |  |  |  |  |  |  |
| SCID |  | 0.42 (s.d.=0.00; Ncohorts=1) |  |  |  |  |  |  | 0.52 (s.d.=0.00; Ncohorts=1) |  |  |  |  |  |  |  |  |  |  |  |
| SITBI |  |  |  | 0.35 (s.d.=0.00; Ncohorts=1) |  |  |  |  |  |  |  |  |  |  |  |  |  |  |  |  |
| SIQ* |  |  |  | 0.50 (s.d.=0.00; Ncohorts=1) | 0.22 (s.d.=0.00; Ncohorts=1) |  |  |  | 0.77 (s.d.=0.00; Ncohorts=1) |  |  |  |  |  |  |  |  |  |  |  |
| DIGS* |  |  |  |  | 0.33 (s.d.=0.00; Ncohorts=1) |  |  |  |  |  |  |  |  |  |  |  |  |  |  |  |
| SSS* |  |  |  |  |  |  |  |  |  |  |  |  |  |  |  | 0.13 (s.d.=0.00; Ncohorts=1) |  |  |  |  |
| MINI* |  |  |  |  |  |  |  |  |  |  |  |  |  |  |  |  | 0.13 (s.d.=0.00; Ncohorts=1) |  |  |  |
| RCADS |  |  |  |  |  |  |  |  |  |  |  |  |  |  |  |  |  | 0.52 (s.d.=0.00; Ncohorts=1) |  | 0.37 (s.d.=0.00; Ncohorts=1) |
| YSR |  |  |  |  |  |  |  |  |  |  |  |  |  |  |  |  |  | 0.48 (s.d.=0.00; Ncohorts=1) | 0.37 (s.d.=0.00; Ncohorts=1) |  |
| CDI |  |  |  |  |  |  |  |  |  |  |  |  |  |  |  |  |  |  | 0.52 (s.d.=0.00; Ncohorts=1) | 0.48 (s.d.=0.00; Ncohorts=1) |

S.d: standard deviation, Ncohorts: number of cohorts

1.	Beck, A. T., Ward. C., Mendelson, M., Mock, J. & Erbaugh J. Beck depression inventory (BDI). Arch Gen Psychiatry. 1961;4:561–571.

2.	Beck A.T., Steer, R.A. & Brown GK. Beck depression inventory-IISan Antonio, TX: Psychological Corporation; 1996.

15.	Beck, A.T., Steer RA. Manual for Beck scale for suicide ideation. San Antonio, TX: Psychological Corporation; 1991.

16.	Poznanski E.O. & Mokros H.B. Children’s depression rating scale, revised (CDRS-R). Los Angeles: Western Psychological Services; 1996.

17.	Cowles Arnette N. Prediction of adolescent suicidality: relative contribution of diagnosis, psychopathy and impulsivity. 2006.

18.	WHO. CIDI-Auto version 1.1: Administrator’s Guide and Reference. 1994.

46.	First MB. Structured clinical interview for DSM-IV Axis I disorders SCID-I: Clinician version, scoresheet1997.

47.	Reynolds W. Suicidal ideation questionnaire (SIQ). Odessa, FL: Psychological Assessment Resources; 1987.

53.	Reynolds W. SIQ, Suicidal Ideation Questionnaire: Professional Manual. 1988.

54.	Beck A, Schuyler D, Herman I. Development of suicidal intent scales. Charles Press Publishers; 1974.

65.	Achenbach T. Manual for the Youth Self Report and 1991 Profile. University of Vermont, Burlington; 1991.

71.	Steer R, Kumar G, Beck A. Self-reported Suicidal Ideation in Adolescent Psychiatric Inpatients. J Consult Clin Psychol. 1993;61.

72.	Valtonen HM, Suominen K, Sokero P, Mantere O, Arvilommi P, Leppämäki S, et al. How suicidal bipolar patients are depends on how suicidal ideation is defined. J Affect Disord. 2009;118:48–54.

73.	Vuorilehto M, Valtonen HM, Melartin T, Sokero P, Suominen K, Isometsä ET. Method of assessment determines prevalence of suicidal ideation among patients with depression. Eur Psychiatry. 2014;29:338–344.

74.	Ballard ED, Luckenbaugh DA, Richards EM, Walls TL, Brutsché NE, Ameli R, et al. Assessing measures of suicidal ideation in clinical trials with a rapid-acting antidepressant. J Psychiatr Res. 2015;68:68–73.

75.	Brown G. A review of suicide assessment measures for intervention research with adults and older adults. 2001.

94.	Lewinsohn P, Rohde P, Seeley J, Baldwin C. Gender Differences in Suicide Attempts From Adolescence to Young Adulthood. J Am Acad Child Adolesc Psychiatry. 2001;40.
